# Unsuppressed HIV infection impairs T cell responses to SARS-CoV-2 infection and abrogates T cell cross-recognition

**DOI:** 10.1101/2022.04.05.22273453

**Authors:** Thandeka Nkosi, Caroline Chasara, Anele Mbatha, Mza Nsimbi, Andrea O Papadopoulos, Tiza L Nguni, Farina Karim, Mohomed Yunus S Moosa, Inbal Gazy, Kondwani Jambo, COMMIT-KZN, Willem Hanekom, Alex Sigal, Zaza M Ndhlovu

## Abstract

HIV infection has been identified as one of the major risk factors for severe COVID-19 disease, but the mechanisms underpinning this susceptability are still unclear. Here, we assessed the impact of HIV infection on the quality and epitope specificity of SARS-CoV-2 T cell responses in the first wave and second wave of the COVID-19 epidemic in South Africa. Flow cytometry was used to measure T cell responses following PBMC stimulation with SARS-CoV-2 peptide pools. Culture expansion was used to determine T cell immunodominance hierarchies and to assess potential SARS-CoV-2 escape from T cell recognition. HIV-seronegative individuals had significantly greater CD4^+^ and CD8^+^ T cell responses against the Spike protein compared to the viremic PLWH. Absolute CD4 count correlated positively with SARS-CoV-2 specific CD4^+^ and CD8^+^ T cell responses (CD4 r= 0.5, p=0.03; CD8 r=0.5, p=0.001), whereas T cell activation was negatively correlated with CD4^+^ T cell responses (CD4 r= −0.7, p=0.04). There was diminished T cell cross-recognition between the two waves, which was more pronounced in individuals with unsuppressed HIV infection. Importantly, we identify four mutations in the Beta variant that resulted in abrogation of T cell recognition. Together, we show that unsuppressed HIV infection markedly impairs T cell responses to SARS-Cov-2 infection and diminishes T cell cross-recognition. These findings may partly explain the increased susceptibility of PLWH to severe COVID-19 and also highlights their vulnerability to emerging SARS-CoV-2 variants of concern.

**One sentence summary:** Unsuppressed HIV infection is associated with muted SARS-CoV-2 T cell responses and poorer recognition of the Beta variant.

## Introduction

Despite measures to contain the spread of SARS-CoV-2 infection, the pandemic is persisting, with a devastating impact on healthcare systems and the world economy (*1*). The research community rapidly mobilized and developed vaccines and therapeutics at unprecedented speed (*2, 3*). COVID-19 vaccines have prevented serious illness and death and have in some cases interrupted chains of transmission at community level (*4*). However, the COVID-19 pandemic remains a major concern in Africa due to dismal vaccine coverage (*5*) and the emergence of variants of concern that may be more transmissible, cause more severe illness, or have the potential to evade immunity from prior infection or vaccination (*6*).

The interaction of HIV-1 infection, common in sub-Saharan Africa, (*7*), with COVID-19 remains understudied. Initial small studies reported that PLWH had similar or better COVID-19 outcomes (*8, 9*). Larger epidemiological studies have demonstrated increased hospitalization and higher rates of COVID-19-related deaths among PLWH compared with HIV negative individuals (*10-13*). Other studies have linked HIV mediated CD4^+^ T cell depletion to suboptimal T cell and humoral immune responses to SARS-CoV-2 (*14*). A recent study showed prolonged shedding of high titre SARS-CoV-2 and emergence of multiple mutations in an individual with advanced HIV and antiretroviral treatment (ART) failure (*15*).

Although B cells have repeatedly been shown to play a pivotal role in immune protection against SARS-CoV-2 infection and antibody responses and are typically used to evaluate immune responses to currently licensed COVID-19 vaccines (*16, 17*), mounting evidence suggest that T cell responses are equally important. For instance, strong SARS-CoV-2-specific T cell responses are associated with milder disease (*14, 18-21*). Moreover, T cell responses can confer protection even in the absence of humoral responses, given that, patients with inherited B cell deficiencies or hematological malignancies are able to fully recover from SARS-CoV-2 infection (*22*). In some instances, COVID-19 disease severity has been attributed to poor SARS-CoV-2-specific CD4^+^ T cell polyfunctionality potential, reduced proliferation capacity and enhanced HLA-DR expression (*14*). Importantly, a recent study identified nonsynonymous mutations in known MHC-1-restricted CD8^+^ T cell epitopes following deep sequencing of SARS-CoV-2 viral isolates from patients, demonstrating the capacity of SARS-CoV-2 to escape from CTL recognition (*23*). Regarding vaccine induced T cell responses, it was recently shown that mRNA vaccines can stimulate Th1 and Th2 CD4^+^ T cell responses that correlate with post-boost CD8^+^ T cell responses and neutralizing antibodies (*24*). The cited examples herein, highlight the need to gain more insight into T cell mediated protection against COVID-19 (*25*).

This study used a cohort of PLWH and HIV-seronegative individuals diagnosed with COVID-19 during the first wave dominated by the wildtype D614G virus (*26*), and the second wave dorminated by the Beta variant. PBMCs were used to determine the impact of HIV infection on SARS-CoV-2 specific T cell responses and to assess T cell cross-recognition. Our data showed impaired SARS-CoV-2 specific T cell responses in individuals with unsuppressed HIV infection and highlighted poor cellular cross-recognition between variants, which was more pronounced than those with unsuppressed HIV. The muted responses in unsuppressed HIV infection may be attributable to low absolute CD4 count and immune activation. Importantly, we identified mutations in the Beta variant that could potentially reduce T cell recognition. Together, these data highlight the need to ensure uninterrupted access to ART for PLWH during the COVID-19 pandemic.

## Results

Study participants were drawn from a longitudinal observational cohort study that enrolled and tracked patients with a positive COVID-19 qPCR test presenting at three hospitals in the greater Durban area. Study participants were recruited into this study based on HIV status and sample availability. They include twenty five participants recruited during the first wave (wild type, wt) of the pandemic in KwaZulu-Natal from June to December 2020 (*27*). Twenty three second wave (Beta variant) participants were recruited from January to June 2021. All study participants where unvaccinated because the COVID-19 vaccine was not readily available in South Africa at the time. Study participants were stratified into three groups, namely HIV-seronegative (HIV neg), People living with HIV (PLWH) with viral load below 50 copies/ml, here termed (suppressed) and PLWH with detectable viral load of ≧1000 copies/ml (viremic). Study participants included HIV-seronegative (HIV neg) (n= 17). PLWH (n=31) were subdivided into suppressed (n=17) and viremic (n=14). The male to female ratio and age distribution were comparable between PLWH and HIV-seronegative groups (Table 1). The median CD4 count for PLWH (suppressed 661 and viremic 301) (p=0.0002, Table 1). Study participants had predominantly mild diseases that did not require supplemental oxygen or ventilation (Table 1).

**Table 1:** Donor characteristics tratified by HIV status

|  | All (n=48) | HIV neg<br>(N=17) | HIV+<br>suppressed<br>(n=17) | HIV+<br>Viremics<br>(N=14) | Statistics |
| --- | --- | --- | --- | --- | --- |
| **Demographics** |  |  |  |  |  |
| Age years, median (IQR) | 40.5(30-51.75) | 45 (27-53.5) | 45(39.5-54) | 31.5 (26.5-42) | 0.036* (KW) |
| Male sex, n(%) | 14 (29.16) | 8 (47.05) | 3 (17.64) | 3 (21.42) | 0.2 (0.82-10) (F) |
| **HIV associated parameters** |  |  |  |  |  |
| HIV viral load<br>Copies/ml |  |  |  | 19969 (2335-43568) |  |
| CD4 cells/ul<br>median (IQR) | 661 (398.5-836.5) | 834.5 (739.3-1029) | 661 (494-789.5) | 301(113.8-568) | 0.0002** (KW) |
| **Disease severity** |  |  |  |  |  |
| Asymptomatic<br>n(%) | 9 (18.75) | 4 (23.52) | 3 (17.64) | 2 (14.28) | 0.6 (0.32-9.53) (F) |
| Mild | 29 (60.42) | 12 (70.59) | 10 (58.82) | 7 (50) | 0.01* (0.13-0.84) (F) |
| Severe/oxygen<br>supplementation | 8 (16.67) | 1 (5.88) | 4 (23.52) | 3 (21.42) | 0.33 (0.48-49.67) (F) |
| Death n(%) | 1 (2.1) | 0 | 0 | 1 (7.1) | 0.46 (F) |

P values calculated by Kruskal-Wallis test for unpaired three groups (KW)
or Fischers exact test (F)

### Unsuppressed HIV infection is associated with altered SARS-CoV-2 specific CD4^+^ and CD8^+^ T cell responses

Immunity to SARS-CoV-2 typically induces robust T cell responses but the impact of HIV infection on these responses has not been fully elucidated (*22, 28, 29*). Thus, we sought to determine the impact of HIV infection on SARS-CoV-2-specific CD4^+^ and CD8^+^ T cell responses. PBMCs were stimulated with PepTivoter 15 mer megapools purchased from Miltenyi Biotec. The pools contained predicated CD4 and CD8 epitopes spanning the entire Spike coding sequence (aa5-1273). Intracellular cytokine staining of peptide stimulated PBMCs was followed by flowcytometric analyses described in the methods section. The samples used for these analyses were collected between two to four weeks after COVID-19 PCR positive diagnosis. Representative flow plots for each group and aggregate data show viremic PLWH had significantly lower frequencies of SARS-CoV-2 specific IFN-γ/TNF-α-produding CD4^+^ T cells compared to suppressed PLWH (p=0.002) and HIV-seronegative individuals (p=0.0006) (Figure 1A). Similarly, viremic PLWH had significantly lower frequencies of SARS-CoV-2 specific IFN-γ/TNF-α-producing CD8^+^ T cells than HIV-seronegative individuals (p=0.02) (Figure 1B). But no significant differences in SARS-CoV-2 specific CD4^+^ T cell or CD8^+^ T cell frequencies was observed between the suppressed PLWH and HIV seronegative individuals (Figure 1A & 1B).

**Figure 1:**
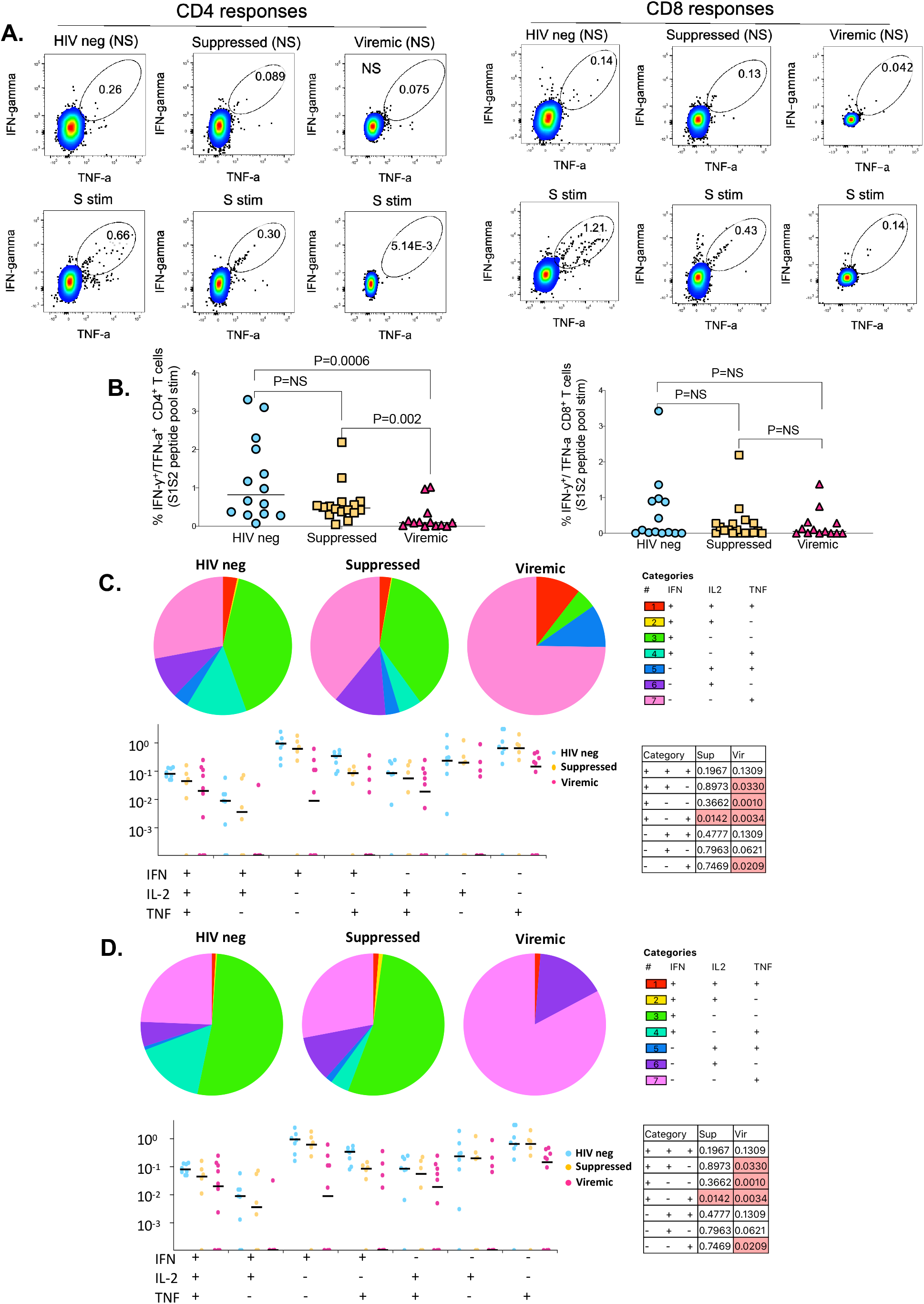
The impact of unsuppressed HIV infection on SARS-CoV-2 specific CD4^+^ and CD8^+^ T cell responses. (**a**) Representative flowplots gated on IFN-γ/ TNF-a dual positive CD4^+^ T cells and aggregate data are shown. (**b**) Representative flowplots gated on IFN-γ/ TNF-a dual positive CD8^+^ T cells and aggregate data are shown (HIV-neg, n=14; suppressed, n=16: viremic, n=13). CD8^+^ T cells producing IFN-γ, TNF-a and IL-2 cells in various combinations are shown. Pie chart and dot plots for by, (**c**) SARS-CoV-2-specific CD8^+^, (**d**) CD4^+^ T cells. Pie chart represents the mean distribution across subjects of mono-functional, bi-functional and poly-functional cytokine producing SARS-CoV-2 specific T cells. Size of each pie segment relates to the frequency of a mono-functional, bi-functional and triple-functional response. Arcs around the pie chart represent the particular cytokine produced. Dot plot represents the frequency of combinations of cytokines produced. Wilcoxon test was done among the dot plots using SPICE software. (Significant p-values are highlighted).

Simultaneous production of cytokines, commonly referred to as polyfunctionality, which is regarded as a measure of the quality of the T cell response, has been shown to correlate with viral control (*30*). Thus, we evaluated the quality of the CD4^+^ and CD8^+^ T cell responses among the groups by enumerating cells co-producing IFN-γ, TNF-α and IL-2. Cells producing all three cytokines were very rare regardless of HIV status (Figure 1C & 1D). The patterns of cytokine production differed in viremic PLWH compared to HIV-seronegative individuals and suppressed PLWH. HIV-seronegative individuals and suppressed PLWH predominantly exhibited IFN-γ responses whereas viremic PLWH predominantly displayed TNF-α responses for both CD8^+^ and CD4^+^ T cells (Figure 1C & 1D). Comparing frequencies of polyfunctional responses between groups confirmed that HIV-seronegative individuals had significantly more dual cytokine producing CD8^+^ and CD4^+^ T cells compared to viremic PLWH (p=0.0330 for CD4 Figure 1C; p=0.0368 for CD8 Figure 1D). Viremic PLWH had significantly lower frequencies of monofunctional IFN-γ producing CD8^+^ and CD4^+^ T cells than suppressed PLWH and HIV seronegative individuals (p=0.0263) (Figure 1C & 1D). Together, these data show that uncontrolled HIV infection lowers the magnitude and alters the quality of SARS-CoV-2 T cells. Importantly, complete plasma HIV suppression preserves the capacity to mount high magnitude dual-functional SARS-CoV-2 specific T cell responses.

### T cell responses against the major SARS-CoV-2 structural proteins

Having observed difference in magnitude and quality of SARS-CoV-2 spike specific T responses, we next measured responses directed against major structural proteins, the nucleocapsid (N) and the membrane (M), again using PepTivoter peptide pools from Miltenyi biotec. Our data show that, although all three major SARS-CoV-2 proteins are targetted, there was a preponderance for T cells to target the S, particularly by HIV-seronegative individuals (Figure 2A & 2B). These data suggest that HIV diminishes Spike specific T cell responses.

**Figure 2:**
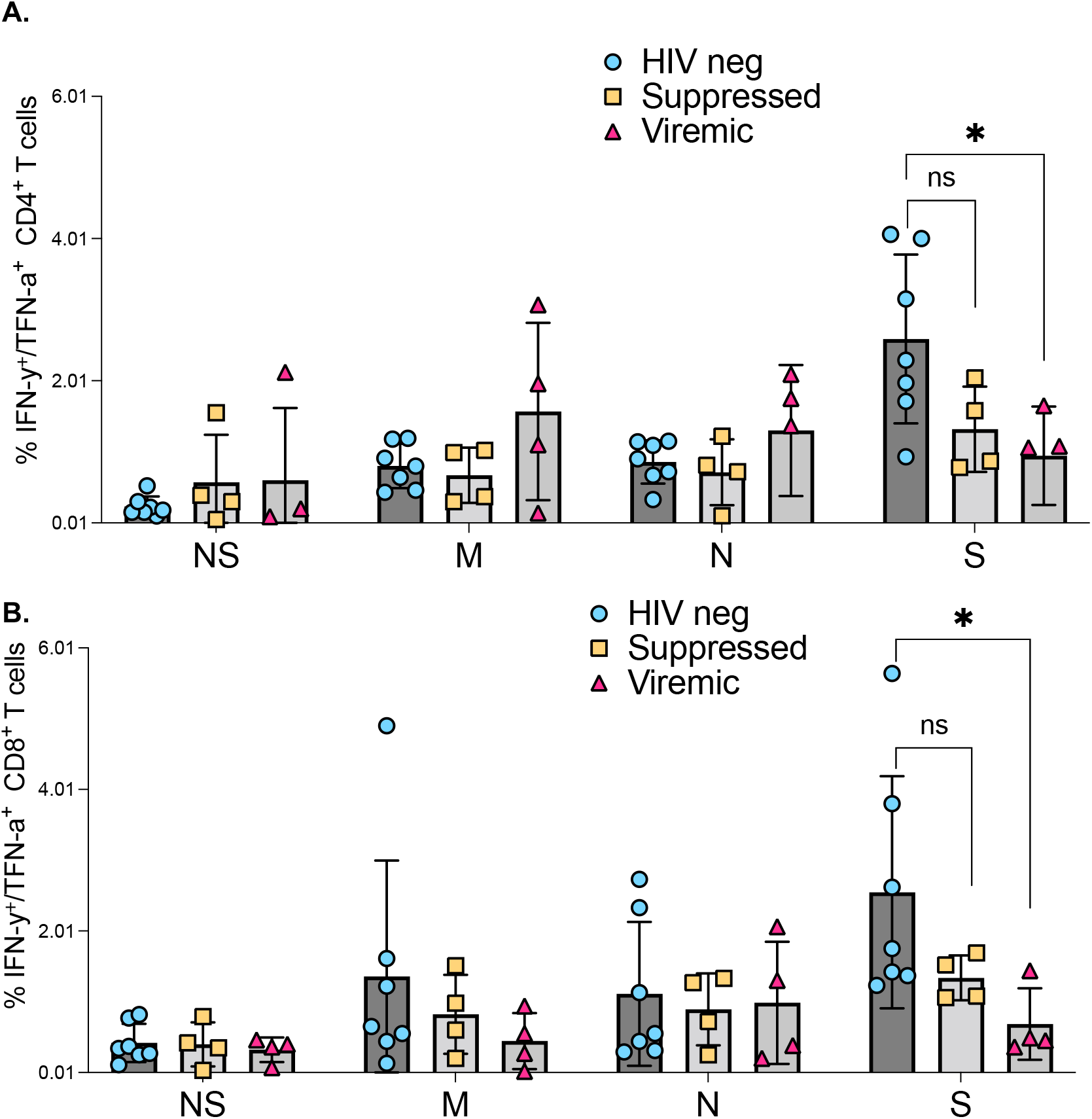
Comparison of SARS-CoV-2 protein targeting by T cell responses among HIV negatives, suppressed and viremic donors: Magnitude of (**a**) CD4^+^ T and (**b**) CD8^+^ T cell responses targetting SARS-CoV-2 proteins among study groups. P-values for differences among the groups are *<0.05; as determined by the Mann-Whitney U test. (GraphPad Prism version 9.3.0)

### Uncontrolled HIV infection abrogates SARS-CoV-2 T cell cross-recognition between wild type D614G and Beta variant

To evaluate the impact of uncontrolled HIV infection on cross reactive T cell responses between wt and the Beta variant, we compared the breadth of responses and the ability to cross-recognize SARS-CoV-2 Beta variant peptides among the three study groups. These studies were conducted using two sets of 15mer overlapping peptides. Set 1 was comprised of 16 wild type (wt) peptides, spanning the receptor binding domain (RBD) and non RBD regions of spike (S) that are known hotspots for mutations (*31*). Set 2 consisted of corresponding peptides that included all the major mutations that define the Beta variant lineage (*32*). A detailed description of the peptides is contained in (Supplementary Table 1).

We first sought to determine cross reactivity of SARS-CoV-2 specific CD4^+^ and CD8^+^ T cells induced following infection with the wild type (D614G, Wave 1) and Beta variant (Wave 2), between each other. We found that wave 1 donors had significantly lower CD8^+^ (p=0.0312) and CD4^+^ T cell responses (p=0.0078) to Beta variant relative to corresponding wt responses (Figure 3A). Wave 2 donors had no significant differences in T cells responses to Beta and wt (Figure 3B). Using a 12 days cultured stimulation assay, we were able to massively expand the magnitude of SARS-CoV-2 specific CD4^+^ and CD8^+^ T cells (Figure 3C, Supplementary Figure 1), and this allowed us to hone in on single peptide responses (Supplementary Table 1). Representative data for a wave 1 donor shows three CD8^+^ and two CD4^+^ positive wt responses (red circles), that did not cross-recognize corresponding Beta variants (blue bars) (Figure 3D). Contrariwise, a representative wave 2 donor had one CD8^+^ and one CD4^+^T cell response to the Beta variant that did not cross-react to the wt version of the peptide (Figure 3E). Intra-donor comparison revealed significantly more CD8^+^ (p=0.0156) and CD4^+^ T cell responses (p=0.0312) to wt peptides compared to the corresponding Beta variant peptides in wave 1 donors (Figure 3F). Conversely, unlike the *ex vivo* data (Figure 3B), wave 2 donors had significantly more CD8^+^ T cell responses to Beta variant peptides relative to wt peptides (p=0.0312), and a trend towards increased CD4^+^ T cells against Beta peptides (p=0.0625), highlighting the increased sensitivity of expanded cells (Figure 3G). Together, these data show poor cross-recognition of wt and Beta variant epitopes.

**Figure 3:**
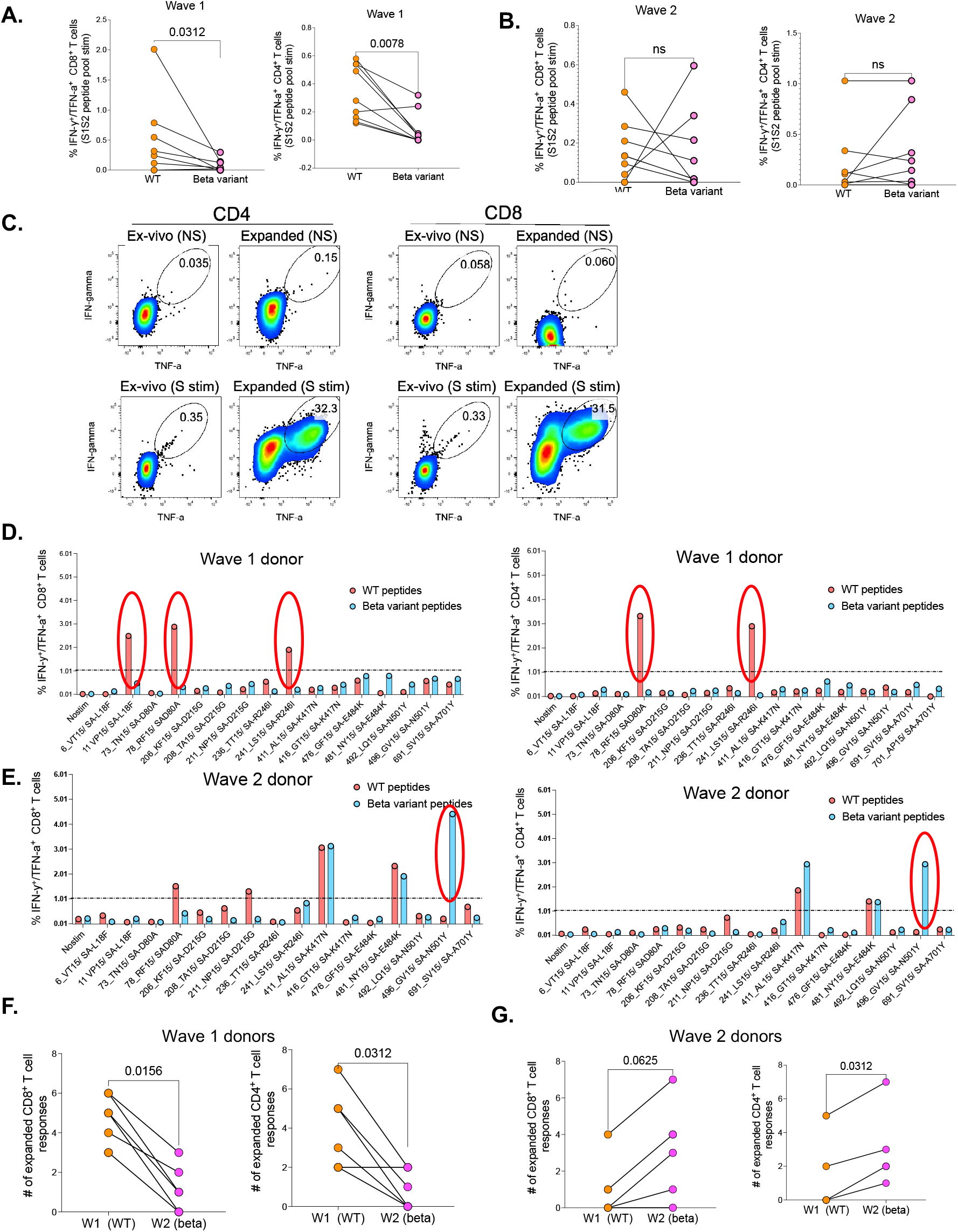
Poor cross-recognition of SARS-CoV-2-specific CD4^+^ and CD8^+^ T cell responses between wt and beta variants in wave 1 and wave 2 COVID-19 participants: *Ex vivo* assessement of T cell cross-recognition between the two waves. (**a**) Intra-donor SARS-CoV-2 specific T cell responses to wt and corresponding Beta variant peptides by wave 1 participants. (**b**) Intra-donor SARS-CoV-2 specific T cell responses to wt and corresponding Beta variant peptides in wave 2 participants. Next, PBMC were expanded for 12 days in the presence of S1S2 SARS-CoV-2 peptide pools and tested against wt and corresponding Beta variants at single peptide level. (**c**) Representative flow plots showing the frequency of SARS-CoV-2 specific CD4^+^ and CD8^+^ T cells before and after cultured expansion. (**d**) T cell responses to single wt (red bars) and corresponding Beta (blue bars) peptide stimulation for a representative donor from wave 1. (**e**) T cell responses to single wt and corresponding Beta peptide stimulation for a representative donor from wave 2. (Positive responses are circled). A response was deemed positive if ≥ 1% or higher. (**f**) Number of wt and corresponding Beta responses for each wave 1 donor. (**g**) Number of wt and corresponding Beta responses for each wave 2 donor. P values calculated using Wilcoxin matched -pairs signed rank T test.

We then assessed the impact of HIV infection on cross recognition of wt and Beta variant epitopes. Representative data for a HIV-seronegative individual from the first wave had 8 wt and 5 Beta variant CD8^+^ T cell responses, one was cross-recognized (circled) (Figure 4A). The same individual had 5 wt and 5 Beta variant CD4^+^ T responses, none was cross-recognized (Figure 4B). Similarly, a representative suppressed wave 1 donor had 5 wt and 2 Beta variant responses one of which was cross recognized (Figure 4C). This same donor had 6 wt and zero Beta variant CD4^+^ T cell responses (Figure 4D). A representative viremic individual had 4 weak wt CD8^+^ T cell responses and 3 borderline CD4 responses, none of which were cross-recognized (Figure 4E & 4F). Summary data showed viremic PLWH had significantly narrow breadth of SARS-CoV-2 specific CD8^+^ (p=0.039) and CD4^+^ T cell responses (p=0.033) compared to suppressed PLWH and HIV seronegative individuals (Figure 4G & 4H). Collectively, these data show that SARS-CoV-2 specific T cell responses in viremic PLWH have limited breadth and subsequently poor cross-recognition potential.

**Figure 4:**
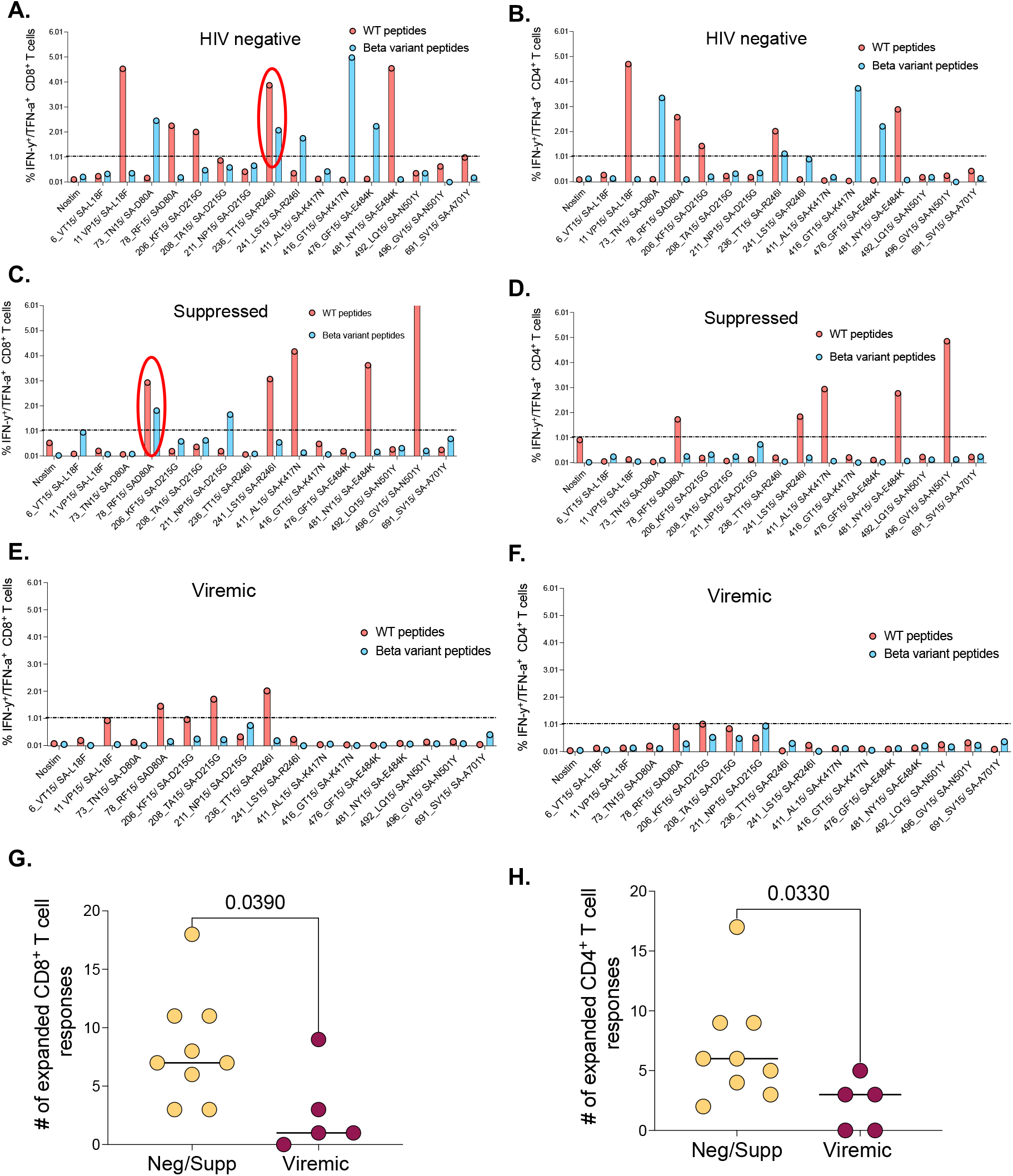
The effects of unsuppressed HIV infection on T cell breadth and ability to cross-recognize the Beta variant: Representative data for a negative donor showing greater, (**a**) CD8^+^ and (**b**) CD4^+^ T cell breadth. A cross-recognized responses between wt and Beta is circled. Representative data for a suppressed donor showing greater, (**c**) CD8^+^ and (**d**) CD4^+^ T cell breadth. A cross-recognized response is circled. Representative data for a viremic donor showing greater, (**c**) CD8^+^ and (**d**) CD4^+^ T cell breadth. (**g**) Aggregate data comparing breath of SARS-CoV-2 specific CD8^+^, and (**h**) CD4^+^ T cell response between HIV negative and suppressed versus viremics. Breadth here is simply the number of positive responses among the individual peptides tested.

### Identification of mutations in the Beta variant that are associated with reduced cross-recognition

Having shown poor T cell cross-recognition of SARS-CoV-2 epitopes between wt and Beta variant, we next sought to identify mutations that might be responsible for the loss of recognition. We combined all the T cell data for the 12 donors used for cultured epitope screening studies. This analysis identified four Beta variant peptides (listed in Table 2) that had significant reduction in CD8^+^ T cell recognition relative to wt peptides (Figure 5A). Three of these peptides were also poorly recognized by CD4^+^ T cells (Figure 5B). The amino acid sequences for wt and corresponding mutations include the E484K mutation, a key Beta variant spike residual change also associated with loss antibody binding (*33*). Together, these data identified mutations in the Beta variant that may abrogate T cell recognition, suggesting that they may be potential T cell escape mutations and warrant further investigation.

**Table 2:**
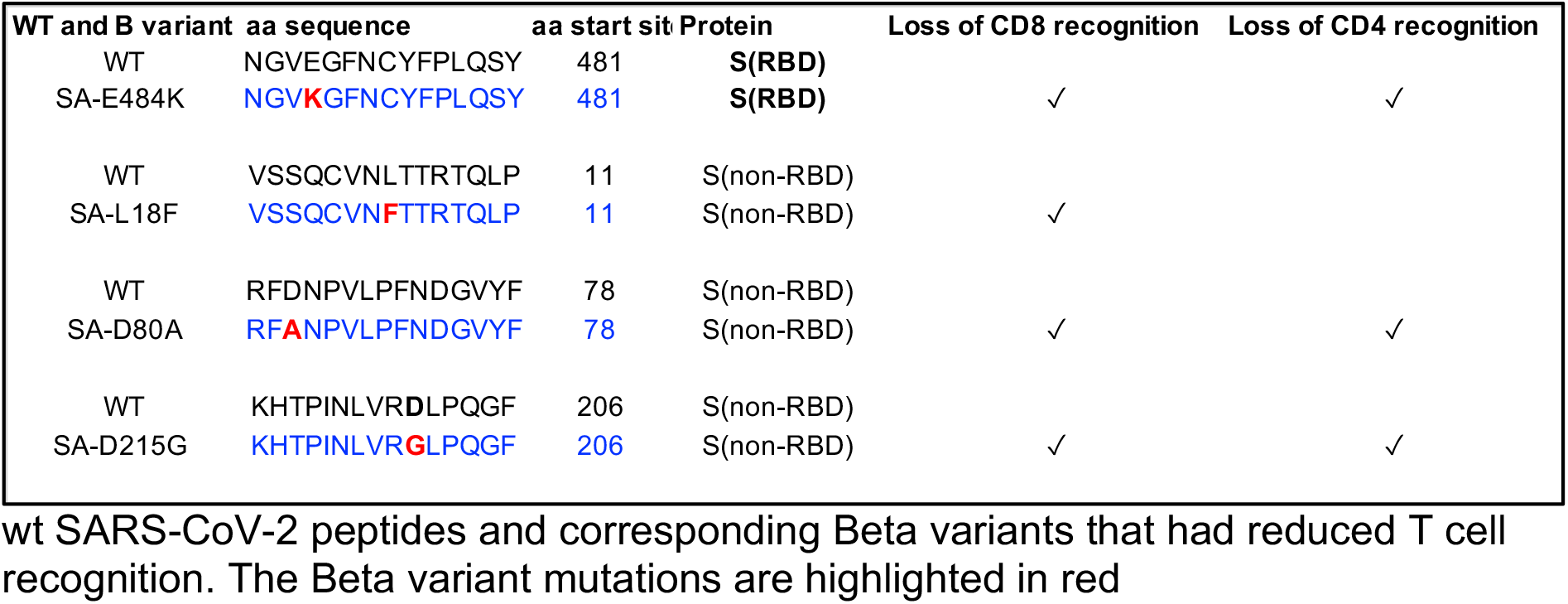
List of 15mer wildtype (wt) and corresponding Beta variant spike peptides sequences

**Figure 5:**
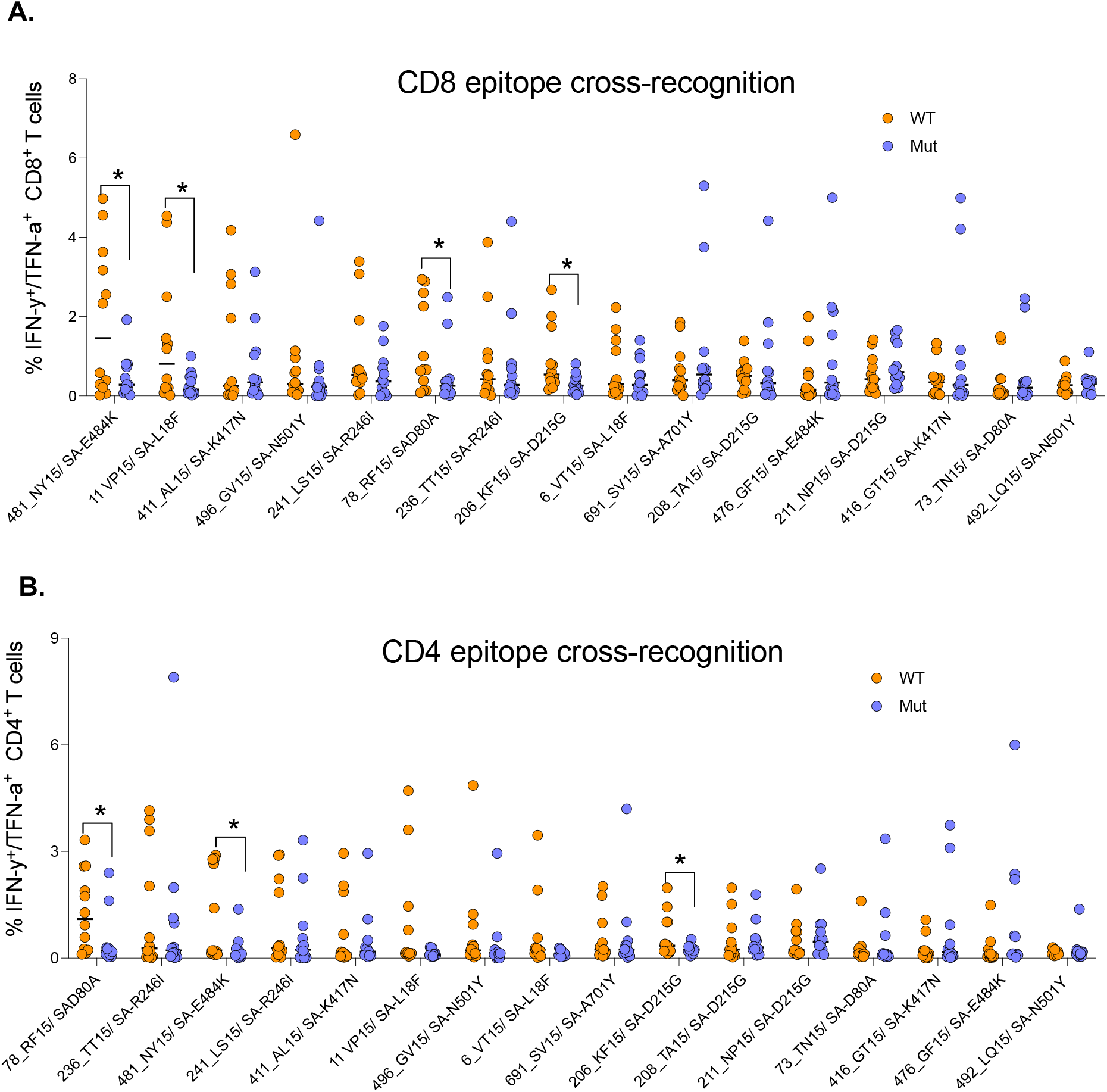
Identification of Beta mutations associated with reduced cross-recognition between wt and Beta variant: (**a**) Side-by-side comparison of SARS-CoV-2 specific CD8^+^ T cell response between wt and Beta. (**b**) Side-by-side comparison of SARS-CoV-2 specific CD4^+^ T cell response between wt. The analysis combined all the 12 participants. P-values calculated by Mann-Whitney U test.

### Immunodominance hierarchy of SARS-CoV-2 CD8^+^ and CD4^+^ T cell responses targeting the spike protein

Virus specific CD8^+^ and CD4^+^ T cells typically target viral epitopes in a distinct hierarchical order (*34, 35*). Identifying SARS-CoV-2 epitopes that are most frequently targeted by T cells is important for the design of vaccines that can induce protective T cell responses. To determine the immunodominance hierarchy of SAR-CoV-2 specific T cell responses targeting the spike protein, OLPs were ranked based on magnitude and frequency of recognition. This analysis revealed the most immunodominant wt peptides targeted by CD8^+^ T cell responses (Figure 6A). The Beta variant resulted in dramatic shift in the immunodominance hierarchy whereby, 3 of 5 most dominant wt CD8^+^ T cell responses (Figure 6A), their Beta variant versions were subdominant (downward arrows) (Figure 6B). Contrariwise, 3 subdominant wt responses were among the most dominant Beta variant responses (upward arrows) (Figure 6B). A similar trend was observed for CD4^+^ T cell responses (Figure 6C & 6D). These data demonstrated a shift in the immunodominant hierarchy between wt and Beta variant responses, which partly explains poor T cell cross-recognition between successive SARS-CoV-2 variants.

**Figure 6:**
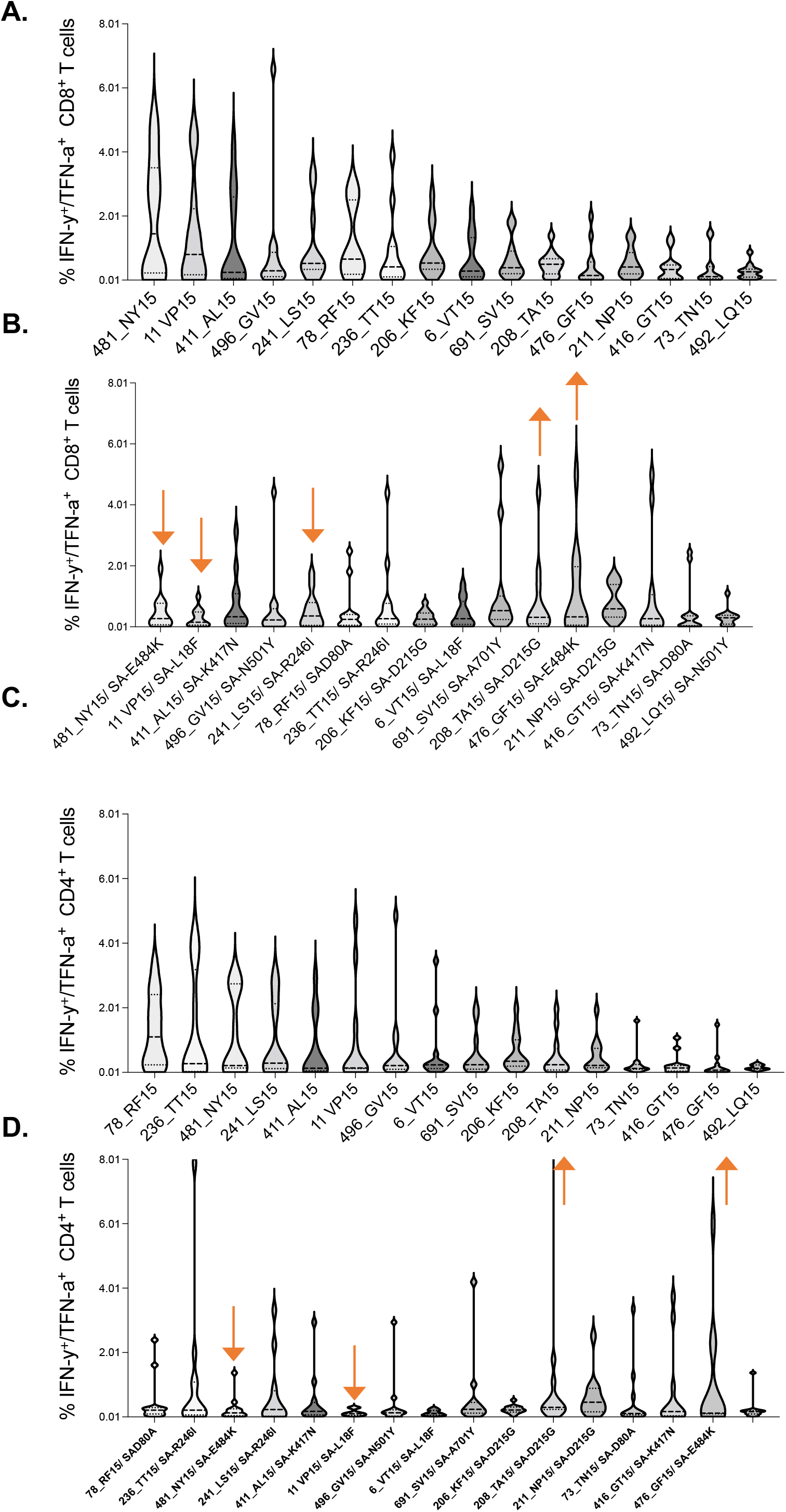
Immunodominance hierarchy of SARS-CoV-2 CD8^+^ and CD4^+^ T cell responses targeting wt and Beta. Immunodminance hierarchy of CD8^+^ T cell responses to, (**a**) wt and (**b**) the corresponding Beta variant peptides. Similarly, Immunodminance hierarchy of CD4^+^ T cell responses to, (**c**) wt and (**d**) the corresponding Beta variant. Arrows indicate responses that changed hierarchical position (among the six most dominant responses) between the two waves. Data arranged in descending order of magnitude of responses to wt peptide stimulation.

### The impact of HIV markers of diseases progression on SARS-CoV-2 specific T cell responses

To gain more insight into why viremic PLWH responded poorly to SARS-CoV-2 infection, we investigated if T cell activation defined here as co-expression of CD38 and HLA-DR, absolute CD4 count and plasma viral load, impacted immune responses (*36*). The proportion of activated (CD38/HLA-DR) SARS-CoV-2 specific CD4^+^ T cells was higher in viremic PLWH compared to suppressed (p=0.02) and HIV seronegative individuals (p=0.002) (Figure 7A). Moreover, proportion of activated (CD38/HLA-DR) SARS-CoV-2 specific CD4^+^ T cells among viremic PLWH negatively correlated with absolute CD4 counts (r=–0.7, p=0.04: Figure 7B), and positively correlated with HIV plasma viral loads (r=0.9, p=0.0004: Figure 7C). Similarly, proportion of activated (CD38/HLA-DR) SARS-CoV-2 specific CD8^+^ T cells were significantly higher in viremic PLWH relative to suppressed PLWH (p=0.04) and HIV seronegative individuals (p=0.0008; Figure 7D). The negative relationship between proportion of activated (CD38/HLA-DR) SARS-CoV-2 specific CD8^+^ T cells and CD4 counts did not reach statistical significance (Figure 7E), but proportion of activated (CD38/HLA-DR) SARS-CoV-2 specific CD8^+^ T cells were positively correlated with HIV plasma viral loads among viremic PLWH (r=0.8, p=0.0006; Figure 7F).

**Figure 7.**
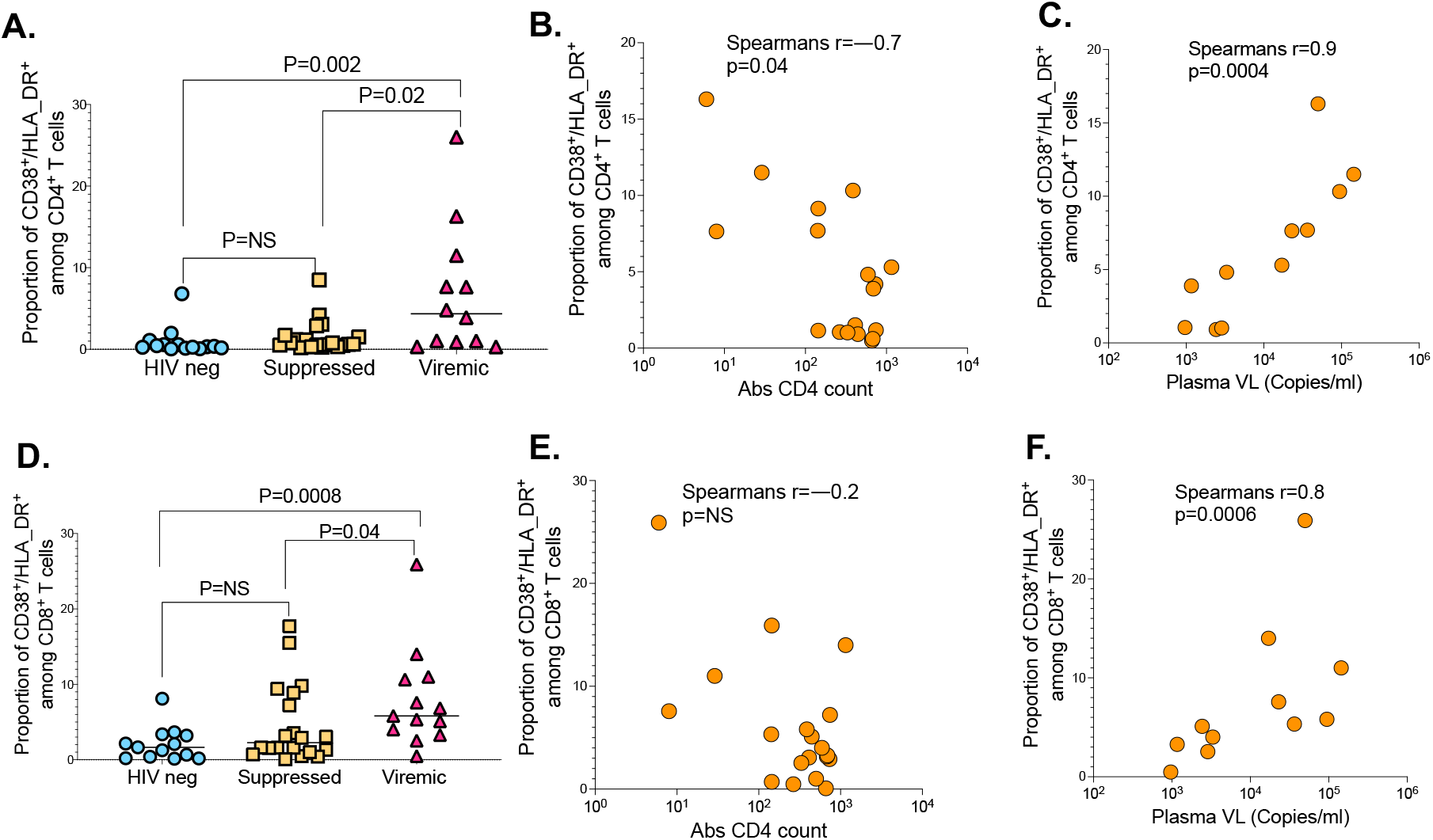
The impact of HIV markers of diseases progression on SARS-CoV-2 T cell immunity. (**a**) CD4^+^ T cell activation graphed based on the frequency of CD38/HLA-DR co-expressing cells. (**b**) Correlation between CD4^+^ T cell activation and absolute CD4 counts. (**c**) Correlation between CD4^+^ T cell activation and HIV plasma viral load. (**d**) CD8^+^ T cell activation measured by CD38/HLA-DR. (**e**) Correlation between CD8^+^ T cell activation and absolute CD4 counts. (**f**) Correlation between CD8^+^ T cell activation and HIV plasma viral load.

Together, these data suggest that hyper immune activation driven by uncontrolled HIV infection impacts SARS-CoV-2 specific CD4^+^ and CD8^+^ T cell responses.

Finally, we interrogated the relationship between SARS-CoV-2 specific responses and disease severity, stratified into asymptomatic, mild and severe disease requiring oxygen supplementation, as previously defined (*27*). We found no significant differences between the magnitude of CD4^+^ or CD8^+^ T cell responses and diseases severity among the groups (Figure S2 A, B). We next, examined sex differences and found no difference in CD4^+^ and CD8^+^T cell responses to SARS-CoV-2 infection (Figure S2 C, D). Age is a risk factor for severe COVID-19 (*5*), thus, we examined the relationship between age and T cell responses. There was a negative relationship between age and magnitude of CD8^+^ T cell responses (CD8 r=-0.6, p=0.002: Figure S2 E), and a similar trend for CD4^+^ T cell responses (CD4 r=-0.3, p=0.15; Figure S2 F). These data show that younger people had greater responses compared to older people whereas, diseases severity and sex did not have discernible effect on SARS-CoV-2 T cell responses.

## Discussion

The greater burden of HIV in sub-Saharan Africa, makes investigating the impact of HIV infection on COVID-19 immunity and disease outcomes critical for bringing the epidemic under control in the region. Recent studies have documented strong cellular responses following SARS-CoV-2 infection and vaccination, but the effects of HIV on SARS-CoV-2 specific T cell responses is not well characterized. Here, we investigated the antigen-specific CD4^+^ and CD8^+^ T cell responses in a cohort of SARS-CoV-2-infected individuals with and without HIV infection. Our results show that unsuppressed HIV infection is associated with reduced cellular responses to SARS-CoV-2 infection. We also show that low absolute CD4 count, and hyper immune activation are associated with diminution of SARS-CoV-2 specific T cell responses. Importantly, we identify spike mutations in the Beta variant that abrogate recognition by memory T cells raised against wt epitopes. Similarly, immune responses targeting Beta variant epitopes poorly cross recognize corresponding wt epitopes. These data reveal the potential for emerging SARS-CoV-2 variants to escape T cell recognition. Importantly, our data highlight the potential for unsuppressed HIV infection to attenuate vaccine induced T cell immunity.

HIV induced immune dysregulation is well documented (*37*). Unsuppressed HIV infection is associated with profound dysfunction of virus-specific T cell immunity partly caused by immune activation (*37, 38*). Our data show that individuals with unsuppressed HIV infection mount weak responses to SARS-CoV-2 infection and poorly recognize SARS-CoV-2 Beta variant mutations. In this study, HIV induced immune defects such as low CD4^+^ T cell counts, higher HIV plasma viral loads and elevated immune activation were invariably associated with diminished SARS-CoV-2 responses. This suggest that HIV induced immune dysregulation negatively impacts the potential to mount robust T cell responses to SARS-CoV-2 infection.

Furthermore, although ART mediated HIV suppression rarely results in complete immune reconstitution (*39*), sustained complete plasma HIV suppression was associated with robust SARS-CoV-2 responses that were mostly similar in magnitude and quality to responses mounted by HIV-seronegative individuals. Given reduced levels of CD38 and HLA-DR dual positive cells and near normal absolute CD4 counts in suppressed individuals, it is reasonable to speculate that reduced immune activation and superior CD4^+^ T helper function were partly responsible for improved immune responses in suppressed individuals.

The emergence of several SARS-CoV-2 variants with mutations in the viral Spike (S) protein such as mutations in the receptor binding domain (RBD), N-terminal domain (NTD), and furin cleavage site region (*40*) continue to fuel the epidemic. These mutations have been shown to directly affect ACE2 receptor binding affinity, infectivity, viral load, and transmissibility (*40-42*). The variants of concern identified since the start of the COVID-19 pandemic include the Alpha (*43*), Beta (*44*), Gamma (*45*), and Delta (*46*) and now the Omicron variant. Most of these have been shown to attenuate neutralization but the impact of these mutations on T cell responses has not been extensively explored (*47*). However, a recent report demonstrating the potential for SARS-CoV-2 to evade cytolytic T lymphocyte (CTL) surveillance, highlight the need for more investigations regarding the potential CTL driven immune pressure to shape emerging variants (*23*). To this end, our study provides new evidence that SARS-CoV-2 has the potential to evade T cell recognition. Moreover, our data suggest that spike mutations in the Beta vatiant that were associated with antibody escape may also escape T cell recognition.

Southern Africa, has had at least three epidemic waves of COVID-19. The first was a mixture of SARS-CoV-2 lineages (with D614G), the second wave was driven by the Beta variant (*48*) and the third by the Delta variant (*49*). The region is currently experiencing the fourth wave dominated by the highly mutated Omicron variant (*50, 51*). Intriguingly, there was some evidence that PLWH in South Africa had increased disease severity in the second wave compared to the first wave(*27*). The precise mechanisms responsible for increased severity are not fully understood, but low CD4^+^ T cell counts and high neutrophil to lymphocyte ratio (NLR) showed strong association with disease severity (*27*). Our data suggest that diminished T cell responses to the Beta variant even in previously exposed individuals may have contributed to severe disease in the second wave.

Although, we repeatedly showed robust *in vitro* T cell expansion following *ex vivo* peptide stimulation but limited expansion against mutant versions of the peptides, there is need to identify optimal peptides that were targeted by CD8^+^ and CD4^+^ T cells in the context of restricting MHC class I and II alleles. SARS-CoV-2 responses are generally very broad (*29*), thus, it is not clear from these studies how loss of T cell cross recognition in Spike affects the overall protective immunity. Furthermore, investigating if the observed poor T cell cross-recognition between wave 1 and wave 2 is generalizable to the Delta and the Omicron variants is clearly warranted. Importantly, our data raises the question of whether CTL selection pressure plays a significant role in shaping emerging variants. This concept should be investigated using larger longitudinal studies with longer durations of follow-up.

In conclusion, we show that uncontrolled HIV infection is associated with low magnitude, reduced polyfunctionality and diminished cross-recognition of SARS-CoV-2 specific CD4^+^ and CD8^+^ T cell responses. Importantly, fully suppressed PLWH had comparable SARS-CoV-2 specific T cell responses with HIV-seronegative individuals. These findings may partly explain high propensity for severe COVID-19 among PLWH and also highlights their vulnerability to emerging SARS-CoV-2 variants of concern, especially those with uncontrolled HIV infection. Hence, there is need to ensure uninterrupted access to ART for PLWH during the COVID-19 pandemic.

## MATERIALS AND METHODS

### Ethical Declaration

The study protocol was approved by the University of KwaZulu-Natal Biomedical Research Ethics Committee (BREC) (approval BREC/00001275/2020). Consenting adult patients (>18 years old) presenting at King Edward VIII, Inkosi Albert Luthuli Central Hospital, and Clairwood Hospital in Durban, South Africa, between 29 July to August November 2021 with PCR confirmed SARS-CoV-2 infection were enrolled into the study.

### Sample collection and laboratory testing

Blood samples used in this study were collected between one to three weeks after COVID-19 PCR positive diagnosis. HIV testing was done using a rapid test and viral load quantification was performed from a 4ml EDTA by a commercial lab (Molecular Diagnostic Services, Durban, South Africa) using the Real Time HIV negative1 viral load test on an Abbott machine. CD4 counts were performed by a commecial lab (Ampath, Durban, South Africa). PLWH were categorised into suppressed and unsuppressed based on viral load measurements of <50 and ≥ 1000 copies/ml respectively, at the time of sample collection.

### T lymphocyte phenotyping

Peripheral blood mononuclear cells (PBMCs) were isolated from blood samples by density gradient method and cryopreserved in liquid nitrogen as previously described (Karim et al., 2020). Frozen PBMCs were thawed, rested, and stimulated for 14 hours at 37 °C, 5% CO_2_ with either staphylococcal enterotoxin B (SEB, 0.5 µg/ml), SARS-CoV-2 wild type peptide pool (8 ug/ml), 501Y.V2 variant peptide pool (4 ug/ml), or the Control Spike peptide pool (Miltenyi, Bergisch Gladbach, Germany, 2 ug/ml). Brefeldin A (Biolegend, California, United States) and CD28/CD49d (BD Biosciences, Franklin Lakes, New Jersey, United States) were also added ahead of the 14-hour incubation at 5 and 1 ug, respectively. The cells were stained with an antibody cocktail containing: Live/Dead fixable aqua dead cell stain, anti-CD3 PE-CF594 (BD), anti-CD4 Brilliant Violet (BV) 650, anti-CD8 BV 786 (BD), anti-CD38 Alexa Fluor (AF) 700 (BD), anti-human leukocyte antigen (HLA) – DR Allophycocyanin (APC) Cy 7 (BD), and anti-programmed cell death protein 1 (PD) BV 421 (BD). After a 20-minute incubation at room temperature, the cells were washed, fixed, and permeabilized using the BD Cytofix/Cytoperm fixation permeabilization kit. Thereafter, the cells were stained for 40 minutes at room temperature with an intracellular antibody cocktail containing: anti-IFN-γ BV 711 (BD), anti-IL-2 PE (BD), and anti-TNF-α PE-Cy 7 (BD). Finally, the cells were washed and acquired on an LSR Fortessa and analysed on FlowJo v10.7.2. Differences between groups were considered to be significant at a *P*-value of <0.05. Statistical analyses were performed using GraphPad Prism 8.0 (GraphPad Software, Inc., San Diego, CA).

### Ex-vivo Cultured expansion of SARS-COV-2 specific T cells

PBMCs at a concentration of 2 million cells per well in a 24-well plate in R10 medium were stimulated with 10 μg/ml of SARS-COV-2 OLPs peptide pools spanning the entire spike protein. The cells were incubated at 37°C in 5% CO_2_. After 2 days, the cells were washed and fresh R10 medium supplemented with 100 U/ml recombinant IL-2 was added. Cultured cells were fed twice weekly with regular medium replenishment. On day 14, the cells were washed three times with fresh R10 medium and rested at 37°C in 5% CO_2_ overnight in fresh R10 medium. On the following day, the cells were simultaneously assessed for their peptide specificity and functional activity by ICS.

### Statistical analyses

All statistical analyses were conducted with GraphPad Prism 9.3.1 (GraphPad Software, La Jolla, California, USA) and *P* values were considered significant if less than 0.05. Specifically, the Mann-Whitney U and Kruskal-Wallis H tests were used for group comparisons. Additional post hoc analyses were performed using the Dunn’s multiple comparisons test. Correlations between variables were defined by the Spearman’s rank correlation test. Categorical data was analysed using the Fisher’s exact test.

## Data Availability

All data produced in the present study are available upon reasonable request to the authors

## Author contributions

A.S, W.H and COMMIT-KZN initiated the study-cohorts. Z.M.N conceived the study and designed the experiments. F.K processed the samples. M.Y, I.G and A.S provided the samples and clinical data. T.N, C.C, A.M, T.J.G and M.S performed the experiments under the supervision of Z.M.N. T.J.N, C.C and Z.M.N and K,J analysed the data. Z.M.N wrote the manuscript. A.P, W.H, A.S and K,J edited the manuscript.

## Acknowledgements

We would like to thank our study participants, the laboratory and clinic staff at Africa Health Research Institute for collecting the samples and compiling the clinical demographic data for the study. We would like to thank Drs Wendy Burgers, Catherine Riou and Robert Wilkinson for desining and providing us the SARS-CoV-2 wt and Beta variant peptides.

This work was funded by; HHMI International research scholar award (Grant #55008743 to ZMN). The AHRI faculty grant (Awarded to ZMN, EW, and HC: LoA R82), The BMGF (Awarded to AS and ZMN (INV-018944), SAMRC (Awarded to WH,# 96838). This work was also partially funded by the Sub-Saharan African Network for TB/HIV Research collaborative award (to ZMN # SANTHE-COL016); Excellence (SANTHE), a DELTAS Africa Initiative (grant # DEL-15-006). The DELTAS Africa Initiative is an independent funding scheme of the African Academy of Sciences (AAS)’s Alliance for Accelerating Excellence in Science in Africa (AESA) and supported by the New Partnership for Africa’s Development Planning and Coordinating Agency (NEPAD Agency) with funding from the Wellcome Trust (grant # 107752/Z/15/Z) and the UK government. The views expressed in this publication are those of the author(s) and not necessarily those of AAS, NEPAD Agency, Wellcome Trust or the UK government.

## Supplementary Material

**Figure S1:**
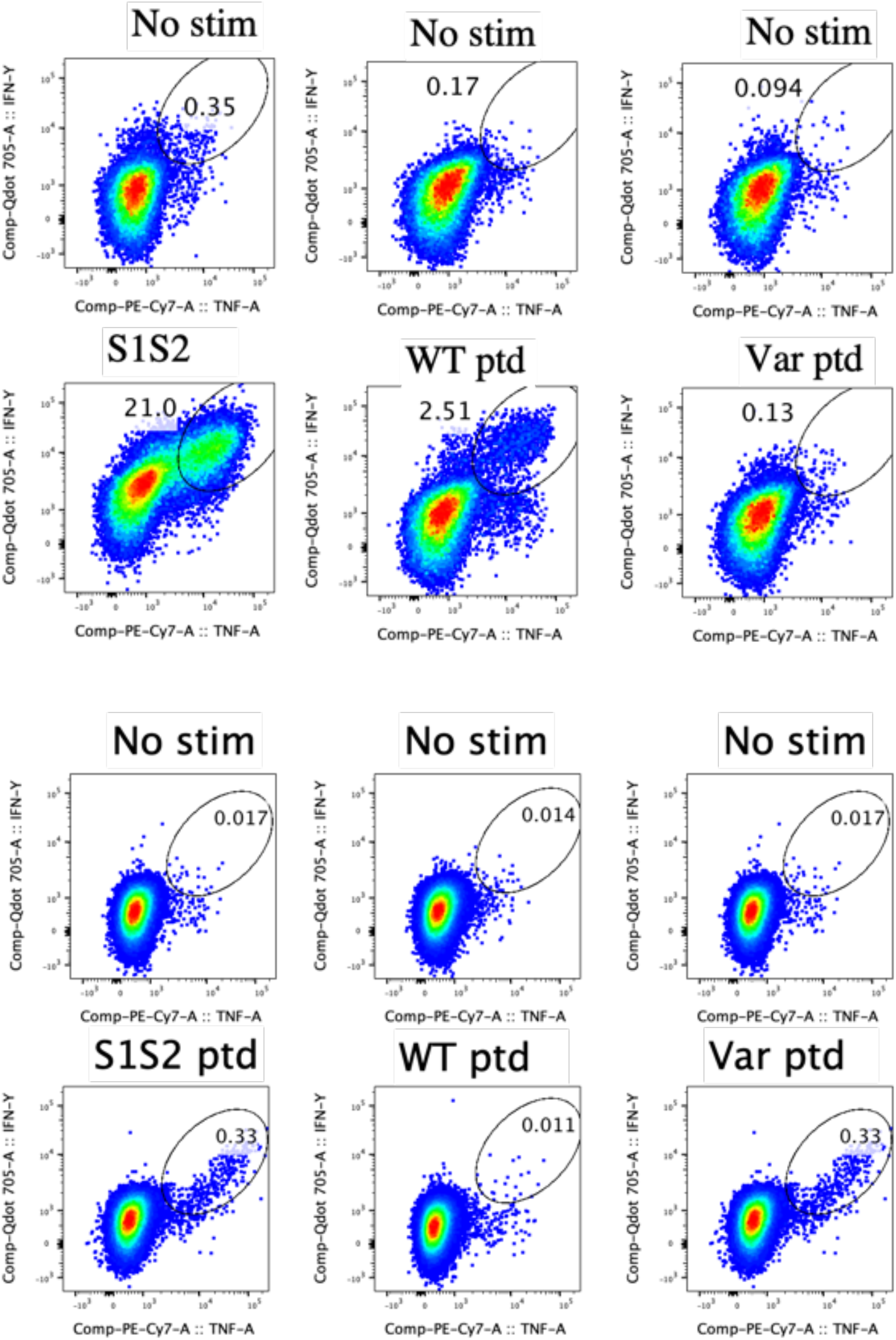
Cross-recognition of SARS-CoV-2 CD4^+^ T cell responses between wt and Beta variants in wave 1 and wave 2 COVID-19 patients: PBMC were expanded for 12 days in the presence of S1S2 SARS-CoV-2 peptide pools. Expanded cells were tested against wt and corresponding Beta variants at single peptide level. (**a**) Intra-donor SARS-CoV-2 specific T cell responses to wt and corresponding Beta variant peptides by wave 1 participants. (**b**) Intra-donor SARS-CoV-2 specific T cell responses to wt and corresponding Beta variant peptides in wave 2 participants.

**Figure S2.**
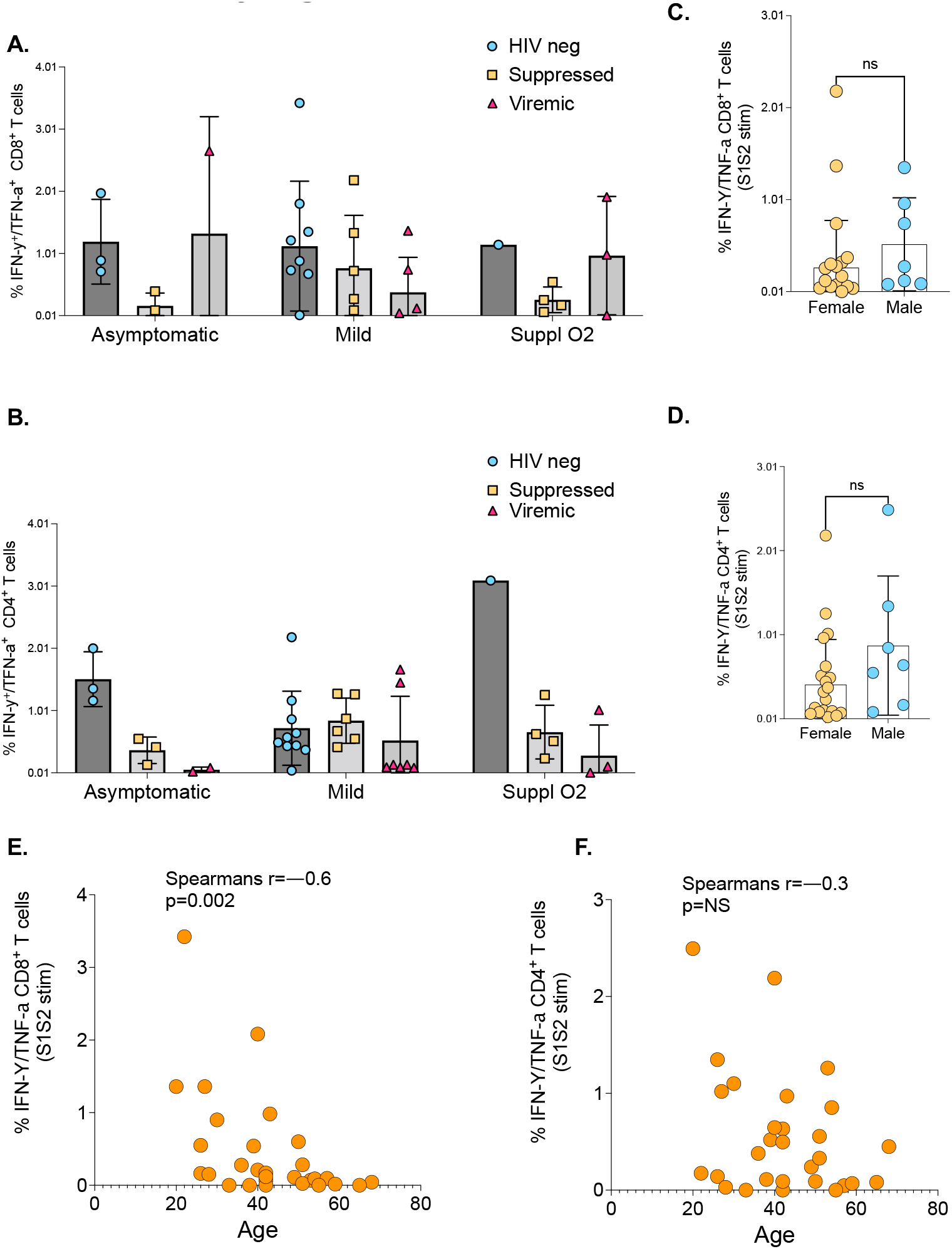
Assessement of the effect of COVID-19 disease severity on, (**a**) SARS-CoV-2 specific CD8^+^, and (**b**) CD4^+^ T cell responses. Disease severity categorised as asymptomatic, mild, and on supplemental oxygen or death. (**c**,**d**) Analysis of SARS-CoV-2 responses based on gender. Correlation between age and SARS-CoV-2 specific, (**e**) CD8^+^T and (**f**) CD4^+^ T cell responses. P-values calculated by Mann-Whitney U test and Pearson correlation test.

**Supplementary table T1:**
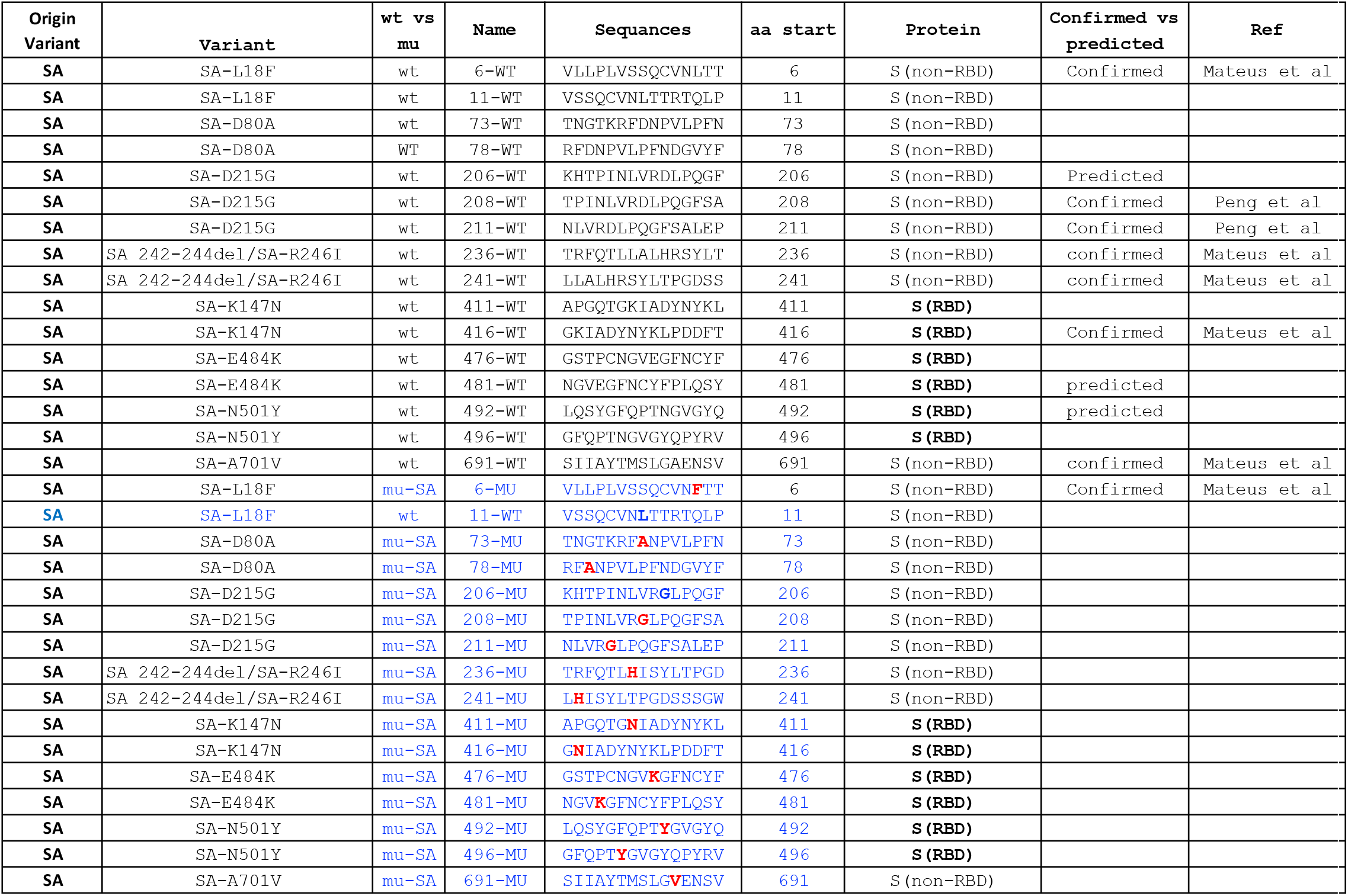
Complete list of 15mer wildtype (wt) and corresponding Beta variant S peptides sequences used for cross-recognition experiments

## References

1. P. Verma et al., A Statistical Analysis of Impact of COVID19 on the Global Economy and Stock Index Returns. SN Comput Sci 2, 27 (2021).

2. F. P. Polack et al., Safety and Efficacy of the BNT162b2 mRNA Covid-19 Vaccine. N Engl J Med 383, 2603–2615 (2020).

3. P. Ball, The lightning-fast quest for COVID vaccines - and what it means for other diseases. Nature 589, 16–18 (2021).

4. G. Kampf, The epidemiological relevance of the COVID-19-vaccinated population is increasing. Lancet Reg Health Eur 11, 100272 (2021).

5. WHO. (2021).

6. V. Shinde et al., Efficacy of NVX-CoV2373 Covid-19 Vaccine against the B.1.351 Variant. N Engl J Med 384, 1899–1909 (2021).

7. H. The Lancet, When pandemics collide. Lancet HIV 7, e301 (2020).

8. L. Calza, I. Bon, M. Borderi, V. Colangeli, P. Viale, No Significant Effect of COVID-19 on Immunological and Virological Parameters in Patients With HIV-1 Infection. J Acquir Immune Defic Syndr 85, e6–e8 (2020).

9. K. W. Lee, S. F. Yap, Y. F. Ngeow, M. S. Lye, COVID-19 in People Living with HIV: A Systematic Review and Meta-Analysis. Int J Environ Res Public Health 18, (2021).

10. M.-A. Davies, HIV and risk of COVID-19 death: a population cohort study from the Western Cape Province, South Africa. medRxiv, 2020.2007.2002.20145185 (2020).

11. A. M. Geretti et al., Outcomes of COVID-19 related hospitalization among people with HIV in the ISARIC WHO Clinical Characterization Protocol (UK): a prospective observational study. Clin Infect Dis, (2020).

12. K. Bhaskaran et al., HIV infection and COVID-19 death: a population-based cohort analysis of UK primary care data and linked national death registrations within the OpenSAFELY platform. Lancet HIV 8, e24–e32 (2021).

13. P. Vizcarra et al., Description of COVID-19 in HIV-infected individuals: a single-centre, prospective cohort. Lancet HIV 7, e554–e564 (2020).

14. C. Riou et al., Profile of SARS-CoV-2-specific CD4 T cell response: Relationship with disease severity and impact of HIV-1 and active <em>Mycobacterium tuberculosis</em> co-infection. medRxiv, 2021.2002.2016.21251838 (2021).

15. F. Karim et al., Persistent SARS-CoV-2 infection and intra-host evolution in association with advanced HIV infection. medRxiv, 2021.2006.2003.21258228 (2021).

16. U. Sahin et al., COVID-19 vaccine BNT162b1 elicits human antibody and TH1 T cell responses. Nature 586, 594–599 (2020).

17. D. S. Khoury et al., Neutralizing antibody levels are highly predictive of immune protection from symptomatic SARS-CoV-2 infection. Nature Medicine 27, 1205–1211 (2021).

18. A. Sette, S. Crotty, Adaptive immunity to SARS-CoV-2 and COVID-19. Cell 184, 861–880 (2021).

19. M. Liao et al., Single-cell landscape of bronchoalveolar immune cells in patients with COVID-19. Nat Med 26, 842–844 (2020).

20. D. Schub et al., High levels of SARS-CoV-2-specific T cells with restricted functionality in severe courses of COVID-19. JCI Insight 5, (2020).

21. C. Rydyznski Moderbacher et al., Antigen-Specific Adaptive Immunity to SARS-CoV-2 in Acute COVID-19 and Associations with Age and Disease Severity. Cell 183, 996–1012 e1019 (2020).

22. E. M. Bange et al., CD8(+) T cells contribute to survival in patients with COVID-19 and hematologic cancer. Nat Med 27, 1280–1289 (2021).

23. B. Agerer et al., SARS-CoV-2 mutations in MHC-I-restricted epitopes evade CD8(+) T cell responses. Sci Immunol 6, (2021).

24. M. M. Painter et al., Rapid induction of antigen-specific CD4(+) T cells is associated with coordinated humoral and cellular immunity to SARS-CoV-2 mRNA vaccination. Immunity 54, 2133–2142 e2133 (2021).

25. D. M. Altmann, R. J. Boyton, SARS-CoV-2 T cell immunity: Specificity, function, durability, and role in protection. Sci Immunol 5, (2020).

26. H. Tegally et al., Sixteen novel lineages of SARS-CoV-2 in South Africa. Nature Medicine 27, 440–446 (2021).

27. F. Karim et al., HIV status alters disease severity and immune cell responses in beta variant SARS-CoV-2 infection wave. Elife 10, (2021).

28. C. Riou et al., Relationship of SARS-CoV-2-specific CD4 response to COVID-19 severity and impact of HIV-1 and tuberculosis coinfection. J Clin Invest 131, (2021).

29. A. Grifoni et al., Targets of T Cell Responses to SARS-CoV-2 Coronavirus in Humans with COVID-19 Disease and Unexposed Individuals. Cell 181, 1489–1501 e1415 (2020).

30. M. R. Betts et al., HIV nonprogressors preferentially maintain highly functional HIV-specific CD8+ T cells. Blood 107, 4781–4789 (2006).

31. H. Tegally et al., Sixteen novel lineages of SARS-CoV-2 in South Africa. Nat Med 27, 440–446 (2021).

32. C. K. Wibmer et al., SARS-CoV-2 501Y.V2 escapes neutralization by South African COVID-19 donor plasma. Nat Med 27, 622–625 (2021).

33. C. K. Wibmer et al., SARS-CoV-2 501Y.V2 escapes neutralization by South African COVID-19 donor plasma. Nature Medicine 27, 622–625 (2021).

34. H. Streeck et al., Human immunodeficiency virus type 1-specific CD8+ T-cell responses during primary infection are major determinants of the viral set point and loss of CD4+ T cells. J Virol 83, 7641–7648 (2009).

35. F. Laher et al., HIV Controllers Exhibit Enhanced Frequencies of Major Histocompatibility Complex Class II Tetramer(+) Gag-Specific CD4(+) T Cells in Chronic Clade C HIV-1 Infection. J Virol 91, (2017).

36. J. Du et al., Persistent High Percentage of HLA-DR(+)CD38(high) CD8(+) T Cells Associated With Immune Disorder and Disease Severity of COVID-19. Front Immunol 12, 735125 (2021).

37. N. R. Klatt, N. Chomont, D. C. Douek, S. G. Deeks, Immune activation and HIV persistence: implications for curative approaches to HIV infection. Immunol Rev 254, 326–342 (2013).

38. Z. M. Ndhlovu et al., Magnitude and Kinetics of CD8+ T Cell Activation during Hyperacute HIV Infection Impact Viral Set Point. Immunity 43, 591–604 (2015).

39. T. J. Henrich et al., HIV-1 persistence following extremely early initiation of antiretroviral therapy (ART) during acute HIV-1 infection: An observational study. PLoS Med 14, e1002417 (2017).

40. A. Tarke et al., Negligible impact of SARS-CoV-2 variants on CD4 (+) and CD8 (+) T cell reactivity in COVID-19 exposed donors and vaccinees. bioRxiv, (2021).

41. A. J. Greaney et al., Comprehensive mapping of mutations in the SARS-CoV-2 receptor-binding domain that affect recognition by polyclonal human plasma antibodies. Cell Host Microbe 29, 463–476 e466 (2021).

42. T. N. Starr et al., Prospective mapping of viral mutations that escape antibodies used to treat COVID-19. Science 371, 850–854 (2021).

43. N. G. Davies et al., Estimated transmissibility and severity of novel SARS-CoV-2 Variant of Concern 202012/01 in England. medRxiv, 2020.2012.2024.20248822 (2021).

44. H. Tegally et al., Emergence and rapid spread of a new severe acute respiratory syndrome-related coronavirus 2 (SARS-CoV-2) lineage with multiple spike mutations in South Africa. medRxiv, 2020.2012.2021.20248640 (2020).

45. C. M. Voloch et al., Genomic characterization of a novel SARS-CoV-2 lineage from Rio de Janeiro, Brazil. medRxiv, 2020.2012.2023.20248598 (2020).

46. S. Mallapaty, India’s massive COVID surge puzzles scientists. Nature 592, 667–668 (2021).

47. C. Riou et al., Escape from recognition of SARS-CoV-2 Beta variant spike epitopes but overall preservation of T cell immunity. Sci Transl Med, eabj6824 (2021).

48. H. Tegally et al., Detection of a SARS-CoV-2 variant of concern in South Africa. Nature 592, 438–443 (2021).

49. E. Callaway, Delta coronavirus variant: scientists brace for impact. Nature, (2021).

50. WHO, Update on Omicron. WHO DASH board. 2021.

51. R. Viana et al., Rapid epidemic expansion of the SARS-CoV-2 Omicron variant in southern Africa. medRxiv, 2021.2012.2019.21268028 (2021).

